# Influence of Nursing Explanation Skills on Incident Occurrence

**DOI:** 10.1101/2023.11.16.23298399

**Authors:** Manabu Fujimoto, Mika Shimamura, Fumiko Yuki

## Abstract

**Objectives:** A routine nursing task is providing explanations to patients, their families, and other healthcare professionals. Inaccurate explanations can adversely affect the quality of healthcare and patient safety. Despite the significance of good explanatory skills in nursing, supporting empirical data are limited. Therefore, this study aimed to develop a psychological scale and investigate the impact of explanatory skills on incidents.

**Methods:** In the preliminary investigation, responses obtained from experienced nurses were analyzed to compile 87 scale items. Study 1 involved an online explanatory skills survey with a sample of 1,000 nursing professionals. Study 2 comprised a field survey involving 159 nursing staff members working in a comprehensive hospital.

**Results:** Nine sub-skills were identified and categorized under two factors: “compassion” and “mental model sharing.” Seven of these sub-skills were found to be shared, and the remaining two were specific to interactions with patients/families or healthcare professionals. Clinical ladder progression was associated with both compassion and mental model sharing, whereas years of practical experience was only related to mental model sharing. Furthermore, compassion was identified as a factor that increased the probability of the occurrence of various incidents through interactional failures. In contrast, mental model sharing enhanced the probability of severe incidents stemming from judgmental failures and minor incidents from conceptual failures.

**Conclusions:** This study developed a psychological scale to measure nursing professionals’ explanation skills in communication with patients, their families, and other medical staff. It elucidated their impact on incident occurrence through miscommunication. The findings need to be practically verified through fieldwork in nursing education.

---

Nursing professionals perform diverse daily tasks.[1][2] Team-based healthcare is common in hospitals, with nursing professionals comprising a significant portion of the medical staff in these teams. They often explain treatment and care by offering decision-making support to patients and their families or ensuring accurate patient handoffs to other medical staff. This highlights the importance of nurses’ explanations to patients, families, and colleagues in healthcare, which has been consistently emphasized in nursing education and practice. For example, “explanation” is an ethically significant concept in healthcare, as shown by its inclusion in the U.S. Belmont Report.[3] Errors in judgment about the communication context or content, neglecting to provide explanations, incorrect information exchange, and shared understanding may cause failures in explanation.[4] These failures are a type of miscommunication. Globally, miscommunication among medical staff affects incident occurrence seriously.[5][6][7] However, the importance of explanations in nursing practice has chiefly been addressed in anecdotal and instructional contexts as communication or informed consent,[8][9][10][11] and there have been few opportunities to discuss this with empirical evidence. Therefore, identifying the explanation skills that nursing professionals need and understanding how these skills impact the quality and safety of nursing treatment and care in nursing communication practices remain crucial challenges.

Given the above background, we accumulated evidence regarding the significance of nursing professionals’ explanations and developed a psychological scale to assess their explanation skills. Furthermore, we elucidated the impact of these skills on patient safety. To achieve our first objective of scale development, we used a scale construction method based on psychological paradigms to identify the skills necessary for nursing professionals when delivering explanations related to treatment and care to patients, their families, and other medical staff. The factors obtained, and their underlying structures revealed key aspects nursing professionals should consider when explaining to patients, families, and colleagues. We expected that the identified factors would be aligned with the definition of “explanation” and encompass elements related to “mental model sharing.” We also expected to identify technical factors related to nursing practice besides cognitive and informational factors. Consistent with many prior studies on nursing communication, this study differentiated explanation recipients into “patients and families” and “medical staff.”[12][13] Consequently, we expected specialized explanation skills to be associated with “informed consent” when communicating with patients and families and with “handoff” when interacting with medical staff.

Several prior studies indicated that many incidents were caused by miscommunication.[5][6][7] Notably, explanation failures comprised a fatal form of miscommunication.[4] Therefore, we postulated that a deficiency in explanation skills may cause miscommunication and increase the probability of incident occurrence through a causal process. Based on the above predictions, our overall hypothesis was that the inadequacy of explanation skills in nursing professionals, as the explainers, leads to miscommunication and triggers incidents. To explore this hypothesis, we constructed models of standard interactions with patients, families, and medical staff and then examined the suitability of these models using multi-group simultaneous path analysis.

## STUDY 1

We collected items of nursing explanation skills using open-ended questionnaires. Next, we identified factors and examined the influence of career quality and quantity on the skills using a web-based survey.

### Methods

#### Preliminary survey

In total, 109 nursing professionals working in hospitals with ≥20 beds and holding clinical ladder positions of II (nurse manager) or higher with over 20 years of experience participated.

We collaborated with an Internet survey company with considerable survey experience and expertise. That company provided access to a panel of medical staff, meaning we could obtain data within a short time frame without the influence of organizational culture. Panel registrants received an introductory email outlining the purpose of the survey and ethical considerations. Those who agreed to participate followed a link in the email to access a dedicated Internet survey site, where they responded and completed the survey by clicking the submission button.

The survey started with items related to participants’ demographic information (Table 1). Next, they were asked to share their opinions based on the instruction, “Please provide your thoughts on behaviors, attitudes, and necessary skills (techniques and abilities) that you consider important when explaining and conveying information about treatment and care to patients/families and other medical staff.”

**Table 1.** Demographic Information.

|  | Preliminary | Study 1 | Study 2 |
| --- | --- | --- | --- |
| Female | 14 | 903 | 138 |
| Male | 95 | 97 | 18 |
| Unanswered | 0 | 0 | 3 |
| Age 20s, y | 0 | 208 | 80 |
| 30s | 0 | 300 | 41 |
| 40s | 68 | 301 | 24 |
| 50s | 36 | 169 | 6 |
| 60s | 5 | 22 | 2 |
| Unanswered | 0 | 0 | 6 |
| Clinical ladder I | 0 | 45 | 33 |
| II | 10 | 206 | 30 |
| III | 35 | 290 | 49 |
| IV | 42 | 159 | 18 |
| V | 22 | 67 | 1 |
| None | 0 | 233 | 28 |
| General Staff | 70 | 843 | 137 |
| Deputy Chief Nurse | 15 | 60 | 11 |
| Chief Nurse | 12 | 40 | 8 |
| Deputy Director of Nursing and Above | 7 | 9 | 0 |
| Non-Regular Staff | 5 | 48 | 3 |
| Nurse | 104 | 926 | 158 |
| Public Health Nurse | 0 | 4 | 0 |
| Midwife | 2 | 24 | 0 |
| Licensed Practical Nurse | 3 | 46 | 1 |
| Hospital (20-199 beds) | 8 | 296 | --- |
| Hospital (200-399 beds) | 76 | 308 | 159 |
| Hospital (<400 beds) | 25 | 396 | --- |

In total, 143 responses were obtained. Item selection was performed in collaboration with all authors, led by the second author, who had extensive experience as a nurse manager. This selection process followed three steps. In step 1, items in each category (“common,” “patients/families,” and “medical staff”) were organized by eliminating duplicate items based on content similarity. Step 2 involved aligning items with identical content across categories. Step 3 comprised modification of item phrasing. Through these processes, a total of 87 items were organized.

#### Main survey

Participants were 1,000 nursing professionals working in hospitals with ≥20 beds. This survey was paper-based and conducted using the same survey company and recruitment method as the initial questionnaire survey. Participants were randomly assigned to either the “patient/family” or the “medical staff” group. In this questionnaire, demographic information was collected first (Table 1), after which participants were asked to rate the 87 items covering nursing explanation skills using a 7- point scale from “very poor” to “very proficient.” If a respondent believed that a particular item did not apply to them (e.g., questions related to explanations to medical staff for the patient/family group), they were instructed to check a box indicating “not applicable” rather than providing a rating.

Exploratory factor analysis was then performed to identify factors. Subsequently, confirmatory factor analysis was used to evaluate the fit of the models for both the patient/family and medical staff groups. Finally, multivariate regression analysis was performed to investigate the impact of career quality and quantity on explanation skills.

#### Ethical considerations

This research was conducted following established ethical guidelines and received approval from Human Research Ethics Committee of Ritsumeikan University (衣笠-人- 2022-106). The preliminary and main surveys were conducted with the assistance of an Internet survey company. Participants in this study were members of a survey panel comprising healthcare professionals. They were informed about this study via email and consented to participate before accessing the survey site.

### Results

#### Item selection

Items related to nursing explanation skills were rated on a 7-point scale, and items participants deemed not applicable to patients/families or medical staff explanations were categorized as “not applicable.” Items marked as not applicable by a certain number of participants were excluded through the following process. On average, 5.45 participants in the patient/family group and 5.33 in the medical staff group marked items as not applicable. Therefore, items marked as not applicable by five or fewer participants were categorized as “applicable (A),” and items marked as not applicable by six or more participants were categorized as “not applicable (NA).” These categories were then combined for patient/family and medical staff interactions. Overall, 49 items in the “applicable-applicable (A-A)” category were shared items, 12 items in the “applicable-not applicable (A-NA)” category were patient/family-specific items, and eight items in the “not applicable-applicable (NA-A)” category were medical staff-specific items. The remaining 18 items in the “not applicable-not applicable (NA-NA)” category were excluded. Data for participants who answered “not applicable” were treated as missing values, resulting in sample sizes for subsequent analyses of 940 participants for shared items (A-A), 467 for patient/family-specific items (A-NA), and 473 for medical staff-specific items (NA-A).

#### Identification of common skills

Using data from Survey 2, an exploratory factor analysis was conducted for the 49 shared items (A-A) (Table 2). This analysis employed the maximum likelihood method, Promax rotation, and the scree criterion. The analysis identified two factors, “compassion” and “mental model sharing.” next, a sub-factor analysis was conducted using the same analysis options (Table 2), which identified seven sub-skills.

**Table 2.** Factor loading and w-weights for A-A items.

|  | **F1** | **F2** | **Sub1** | **Sub2** | **Sub3** | **Sub4** |
| --- | --- | --- | --- | --- | --- | --- |
| 47 Speaking in a voice that is easy for the other person to hear | **.591** | .186 | **.772** | .091 | .018 | -.067 |
| 58 Making eye contact with the other person | **.581** | .101 | **.733** | -.023 | .073 | -.053 |
| 66 Starting the conversation with a greeting | **.726** | .021 | **.714** | -.040 | .125 | .015 |
| 72 Adjusting the speaking speed appropriately | **.613** | .219 | **.471** | .149 | -.120 | .366 |
| 67 Building a good relationship with the other person beforehand | **.680** | .138 | **.458** | .335 | .055 | .025 |
| 69 Explaining while observing the other person's reactions | **.607** | .278 | **.442** | .324 | -.019 | .066 |
| 52 Inferring the other person's emotions from their expressions | **.608** | .157 | .065 | **.693** | .103 | -.046 |
| 86 Explaining according to the other person's personality and character | **.552** | .317 | .108 | **.628** | -.075 | .241 |
| 48 Considering the other person's position and individuality | **.664** | .200 | .356 | **.465** | .107 | -.010 |
| 23 Confirming the other person's way of speaking and the content | **.545** | .300 | .067 | **.432** | .243 | .136 |
| 19 Respecting the other person | **.840** | -.062 | .100 | -.139 | **.855** | .096 |
| 21 Adopting the other person's perspective | **.730** | .084 | -.134 | .320 | **.637** | -.017 |
| 11 Explaining politely | **.511** | .294 | .295 | .054 | **.532** | -.032 |
| 44 Empathizing with the other person's feelings | **.912** | -.225 | .123 | .135 | **.500** | .162 |
| 14 Repeating the explanation multiple times | **.378** | .352 | .072 | .163 | **.442** | .076 |
| 87 Not imposing your thoughts | **.769** | -.026 | -.080 | .160 | .009 | **.753** |
| 73 Not denying the other person's claims | **.865** | -.092 | .184 | -.083 | .082 | **.707** |
| 24 Not becoming emotional | **.452** | .114 | -.168 | .032 | .277 | **.491** |
| 63 Actively listening to the other person | **.957** | -.214 | .251 | -.054 | .274 | **.386** |
|  | w |  | .912 | .894 | .909 | .841 |
| 65 Confirming how well the other person understands after explaining | .437 | **.447** | **.886** | -.060 | .059 |  |
| 46 Confirming how the other person perceives the current situation | .374 | **.508** | **.699** | .128 | .063 |  |
| 78 Providing accurate information to the other person | .355 | **.514** | **.638** | .204 | .025 |  |
| 41 Considering how much detail to provide | .207 | **.648** | **.578** | .136 | .178 |  |
| 83 Checking for questions at the end of the explanation | .217 | **.567** | **.550** | .272 | -.026 |  |
| 22 Conveying facts without inserting subjectivity | .235 | **.542** | .186 | **.643** | -.037 |  |
| 25 Providing explanations based on evidence | -.093 | **.894** | .053 | **.594** | .240 |  |
| 54 Concisely explaining the basis and policies | .054 | **.780** | .238 | **.548** | .091 |  |
| 32 Providing information gradually when there is a lot of it | .131 | **.648** | .334 | **.524** | -.046 |  |
| 15 Conveying information in the order of the situation, background, thoughts, and proposals | -.020 | **.806** | .130 | **.412** | .316 |  |
| 2 Considering the order of priority in the explanation | -.238 | **.899** | -.097 | .134 | **.754** |  |
| 1 Creating an environment conducive to conversation | .207 | **.458** | .204 | -.199 | **.712** |  |
| 9 Explaining with an analysis of the progress and situation | -.214 | **.963** | -.050 | .285 | **.639** |  |
| 10 Adapting communication to the individuality of the other person | .326 | **.490** | .169 | .070 | **.609** |  |
|  | w |  | .917 | .899 | .866 |  |
| The bold items represent factor loadings for the factors with the highest loadings. |  |  |  |  |  |  |

#### Identification of specific skills and validation of the model

Another exploratory factor analysis was conducted using the same analysis options to identify specific skills related to explanations for patients/families and medical staff (Table 3). The analysis of the 12 items for patients/families (A-NA) identified “agreement” as a sub-skill, and the analysis of the eight items for medical staff (NA-A) identified “handoff” as a sub-skill.

**Table 3.** Factor loading and w-weights for A-NA and NA-A items.

|  |  |
| --- | --- |
| 84 Align the image with the other party if it deviates. | **.849** |
| 13 Convey sufficient information to reach an agreement. | **.843** |
| 62 Ask the same question differently. | **.805** |
| 49 Confirm what state the other party wants to be in. | **.794** |
| 77 Encourage open expression of requests. | **.764** |
| 26 Explain using materials. | **.763** |
|  | w .916 |
| 31 Utilize notes for evident explanations. | **.795** |
| 27 Maintain records for information sharing. | **.788** |
| 88 Ensure effective communication through reports and messaging. | **.771** |
| 81 Confirm the understanding of specialized matters. | **.742** |
| 8 Thoroughly implement reporting, contacting, and consulting practices. | **.712** |
|  | w .873 |
| The bold items represent factor loadings for the factors with the highest loadings. |  |

Scale scores for each sub-skill were calculated to confirm whether the two identified specific skills were included in the compassion or mental model sharing factors. An exploratory factor analysis was conducted for the seven shared skills and one specific skill (“agreement” for the patient/family group and “handoff” for the medical staff group) to verify their inclusion. The results showed that both specific skills loaded onto mental model sharing.

Therefore, a hierarchical factor analysis model (Figure 1) was validated for the patient/family and medical staff groups, which included the specific skills (agreement/handoff) within the sub-skills. Confirmatory factor analysis showed acceptable fit indices for both the patient/family group (goodness of fit index [GFI]=0.975, comparative fit index [CFI]=0.992, root mean square error of approximation [RMSEA]=0.087) and the medical staff group (GFI=0.977, CFI=.992, RMSEA=0.083).

**FIGURE 1.**
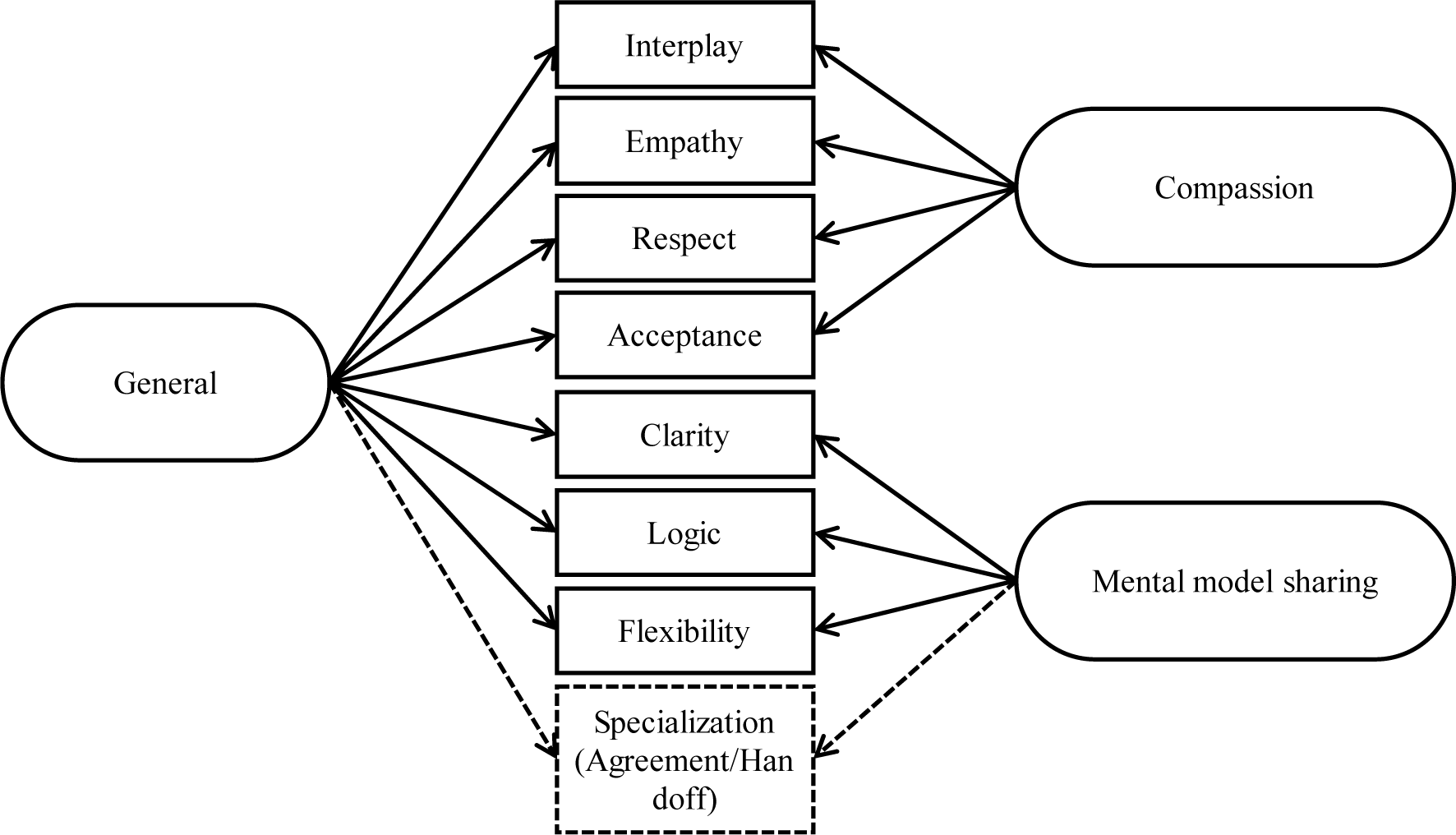
Hierarchical Factor Model of Nursing Professionals’ Explanation skills. Squares denote the observed variables and ovals denote the latent variables. Solid lines represent factors specific to A-A items, and dotted lines represent factors specific to A-NA and NA-A items.

#### Verification of the impact of a nursing career on explanation skills

Multivariate regression analysis was conducted to examine the impact of a nursing career on explanation skills. The explanatory variables included the explanation target (i.e., patient/family group or medical staff group), years of practical experience, clinical ladder, and interaction terms. The outcome variables were the scale scores for the explanation skills sub-skills. The overall model was significant (*R^2^*=.220, Pillai’s trace=.237, *F*(56,6489)=4.06***). Among the explanatory variables, years of practical experience (*F*=11.54***) and clinical ladder (*F*=6.17***) were significant. The years of practical experience showed a significant relationship with the shared skills in the mental model sharing factor (clarity*, logic**, flexibility**). The clinical ladder significantly impacted all explanation skills (acceptance and seven sub-skills).

### Discussion

This study elucidated the hierarchical factor structure for nursing explanation skills. In addition to identifying mental model sharing as anticipated based on the conceptual definition of explanation, we identified compassion as an upper-level factor. Compassion represented a socioemotional and relationship-maintaining skill that involved consideration for others. Conversely, mental model sharing encompassed cognitive and problem-solving skills to facilitate communication and understanding with others. Various concepts in interpersonal psychology are typically categorized into cognitive (task) and socioemotional (socioemotional) aspects. This dichotomy was also observed in nursing explanation skills. The first factor (compassion) corresponded to socioemotional factors, whereas the second (mental model sharing) matched cognitive factors.

The analysis of the impact of nursing careers revealed that accumulating quantitative career experience could enhance the techniques in mental model sharing (clarity, logic, flexibility) could be enhanced by accumulating quantitative career experience. However, to acquire and practice mental model sharing and compassion, it was evident that it should enhance the quality of a nurse’s career in line with the clinical ladder criteria aligned with the organization’s nursing education philosophy. The two significant skills were subdivided into nine sub-skills related to the sub-factors. Seven sub-skills were common to patient/family and medical staff interactions. Specific sub-skills for the target in the mental model sharing construct included “agreement” for patient/family and “handoff” for medical staff. These target-specific sub-skills reflected differences between patients/family and medical staff regarding the purpose of communication and content to be conveyed and variations in specialized knowledge related to healthcare. The other seven common sub-skills were considered essential elements for the act of explanation in general and not limited to nursing.

## STUDY 2

This study involved a hospital survey to confirm the reliability and validity of the developed scale.

Next, we examined the impact of nursing explanation skills on incidents.

### Methods

#### Participants

173 nursing professionals working in a single comprehensive hospital with ≥350 beds participated. Of these, 159 responses were considered valid (85 from the patient/family group and 74 from the medical staff group).

#### Survey procedure

The survey was conducted using Google Forms. Participants accessed a website that displayed the same content as the guidebook. Only those who agreed to participate in this study could access either the “Patient/Family Questionnaire” or the “Medical Staff Questionnaire” through a link on a single page of the Google Form. This was presented randomly via JavaScript.

#### Survey content

Participants provided demographic information (Table 1). Next, two nursing explanatory skill scales were completed, one for patient/family (shared skills and agreement) and the other for medical staff (shared skills and handoff). As in Study 1, participants rated these skills on a seven-point scale from “very poor” to “very proficient.” The ENDCOREs scale^(14(15^ was used to measure general communication skills and confirm the criterion-related validity of our developed scale. This scale measures six skills: self-control, expressivity, sensitivity, assertiveness, responsiveness, and regulation, giving 24 items. Ratings for these factors were collected using a seven-point scale (“very poor” to “very proficient”). Finally, participants completed a questionnaire related to miscommunication and patient outcomes.^(4^ For miscommunication, six “situational failure type” items and two “conceptual failure type” items were rated using a seven-point scale (from “never” to “always”). Four items related to patient outcomes were assessed using a seven-point interval scale (from “none” to “always”).

#### Analytical methods

First, we calculated the omega coefficient to obtain reliability. Next, a correlation analysis was performed for the nursing explanatory skill scale and ENDCOREs factors to verify criterion-related validity. Finally, a multi-group simultaneous path was used to validate the hypothetical model that nursing explanation skills cause incidents through miscommunication.

#### Ethical considerations

This study was conducted after obtaining approval from Human Research Ethics Committee of Ritsumeikan University (衣笠-人-2022-106). Consent was also obtained from the hospital administrators, and a document requesting cooperation in the survey was distributed to hospital nurses. This document outlined the purpose and methodology of the study, emphasized voluntary participation, and assured participants that the data would be statistically processed and individuals would not be identifiable. Those who understood and agreed to cooperate accessed the survey via Google Forms.

### Results

#### Reliability and validity of explanation skills

The reliability of explanation skills (as indicated by the omega coefficients) and the correlation between explanation and communication skills were verified (Table 4). Explanation and communication skills positively correlated (*r*=.249–.730). In addition, the clear, logical, and flexible aspects of mental model sharing demonstrated strong positive correlations with expressive ability and self-assertion in the expressive system. The four compassion sub-skills and the “specific” category exhibited strong positive correlations with the interpretative ability and acceptance of others in the responsive system and self-regulation and relationship management in the management system.

**Table 4.** Correlation Coefficients Between Explanation Skills and Communication Skills.

|  | Self Control | Expressivity | Sensitivity | Assertiveness | Responsiveness | Regulation |
| --- | --- | --- | --- | --- | --- | --- |
| Interplay | .493 ** | .393 ** | .588 ** | .529 ** | .632 ** | .624 ** |
| Empathy | .582 ** | .384 ** | .730 ** | .510 ** | .613 ** | .660 ** |
| Respect | .572 ** | .405 ** | .607 ** | .506 ** | .659 ** | .662 ** |
| Acceptance | .627 ** | .249 ** | .499 ** | .332 ** | .676 ** | .589 ** |
| Clarity | .541 ** | .456 ** | .623 ** | .586 ** | .549 ** | .658 ** |
| Logic | .528 ** | .473 ** | .560 ** | .577 ** | .434 ** | .591 ** |
| Flexibility | .589 ** | .413 ** | .613 ** | .539 ** | .529 ** | .652 ** |
| Specialization | .539 ** | .420 ** | .617 ** | .527 ** | .579 ** | .609 ** |

#### Hypothesis model setup

Based on the hypothesis that the lack of explanation skills in nursing professionals (who function as communicators) leads to miscommunication and triggers incidents, we developed a conceptual model to illustrate specific relationships between variables obtained in this study (Figure 2). Many concepts in interpersonal psychology tend to be dichotomized into socioemotional and cognitive variables.[16][17] This study identified socioemotional compassion and cognitive mental model sharing as representing nursing professionals’ explanation skills. Therefore, the paths between variables in the hypothesis model were set along socioemotional and cognitive routes.

**FIGURE 2.**
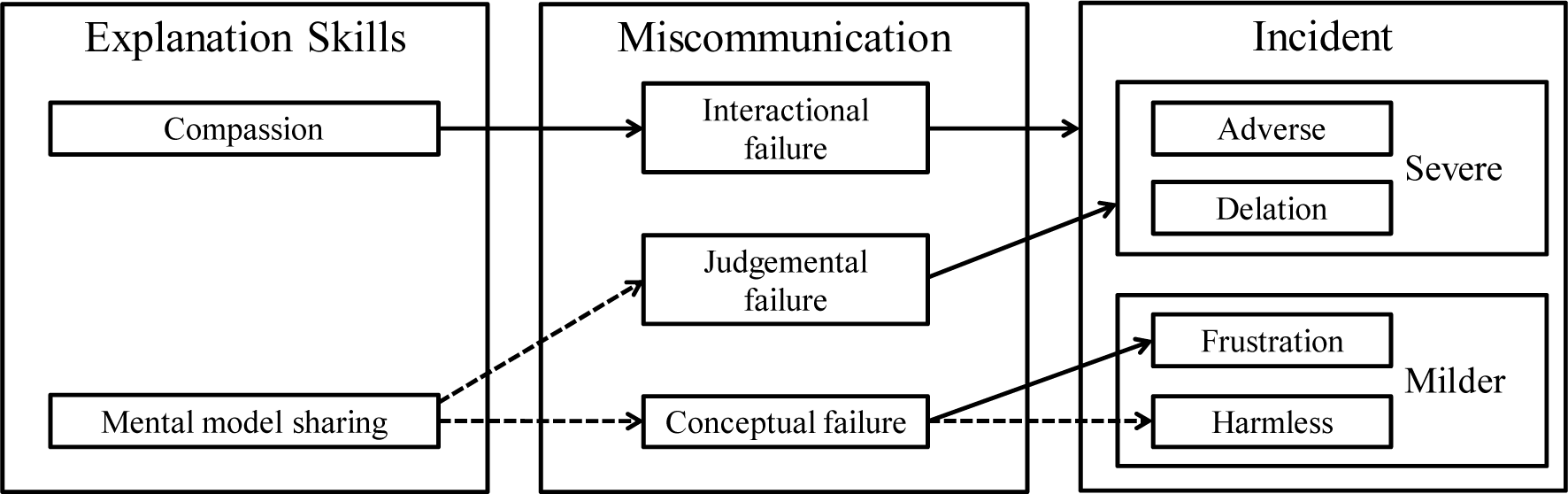
Hypothetical Model of Incident Occurrence. Dotted lines represent stronger associations with explanations for healthcare professionals rather than for patients/families.

In the first part of the hypothesis regarding the “impact of explanation skills on miscommunication,” compassion, a skill characterized by its socioemotional nature, was set to have a path with the interactional failure type, where communication is neglected. Conversely, mental model sharing, a skill characterized by its cognitive nature, had paths with the judgmental failure type related to misjudgments in situations, contexts, and information and the conceptual failure type associated with insufficient information exchange and shared understanding.

Next, regarding the part of the hypothesis concerning the influence of miscommunication on incidents, a path was set between the interactional failure type and all incident indicators because this was the communication type in the socioemotional route. As miscommunication in the cognitive route was divided into judgmental and conceptual failure types, we set a path based on the severity of the incident. The judgmental failure type, in which the patient misjudges the situation, or the information conveyed, seriously impacts treatment and care.[18] Therefore, another path from judgmental failure type to “adverse” and “delay” was established. The conceptual failure type encompasses miscommunication, where information exchange and unification of intention were attempted but inadequate. For example, the issue of informed consent causes unnecessary frustration to patients and their families.[19] In addition, a lack of unification among healthcare providers leads to near-misses[20]. Therefore, a path from conceptual failure type to “frustration” and “harmless” was established.

#### Hypothesis model validation

The hypothesis model for the patient and medical staff groups was jointly validated using a maximum likelihood multi-group path analysis (Figure 2). First, an unconstrained model was verified and a good model fit was obtained (minimum discrepancy function divided by degrees of freedom [CMIN/DF]=1.30, *p*=.129; root mean square residual [RMR]=0.065, GFI=0.950, CFI=0.987, RMSEA=0.043, Akaike information criterion [AIC]=158.86). Therefore, configure invariance was confirmed. Next, we examined differences in parameter estimates between the groups. The results indicated significant differences in the path coefficients from mental model sharing to judgmental failure type (patients: −0.03, medical staff: −0.38; *z*=2.33) and conceptual failure type (patients: 0.00, medical staff: −0.46; *z*=3.20), as well as the error variances between delay and frustration (patients: 0.38, medical staff: 0.76; *z*=−2.80), all of which exceeded the absolute value of 1.96.

Therefore, we tested partially constrained equivalence models, resulting in an acceptable model fit (CMIN/DF=1.88, *p*=.001; RMR=0.144, GFI=0.920, CFI=0.956, RMSEA=0.043, AIC=175.77).

Furthermore, fully constrained equivalence models were tested for all paths and correlations, which showed an acceptable model fit (CMIN/DF=1.47, *p*=.018; RMR=0.141; GFI=0.915, CFI=0.967, RMSEA=0.055, AIC=154.67). Therefore, measurement invariance was confirmed. Finally, the three models were compared. There were significant differences between the unconstrained model and both the partially constrained equivalence model (*χ^2^*(4)=24.91, *p*<.001) and the fully constrained equivalence model (*χ^2^*(18)=31.81, *p*=.023). However, there was no significant difference between the partially and fully constrained equivalence models (*χ^2^*(14)=6.91, *p*=.938). After comprehensively comparing the model fit indices of the unconstrained model and the partially/fully constrained equivalence models, an unconstrained model was adopted. This suggested that the impact structure was common between patients/families and medical staff, but there were differences in the strengths and weaknesses of the influences among some elements.

### Discussion

The scale developed in Study 1 was confirmed to be reliable and valid. We named this scale the “SNES” (Scale of Nursing Explanation Skills). Regarding the influence of nursing explanation skills on the occurrence of incidents, we found that in both patient/family and medical staff interactions, nursing professionals who struggled with compassion tended to commit more interactional failures by avoiding interactions with patients/families and other medical staff. Through these transaction failings, these professionals contributed to various incidents, including adverse events, treatment and care delays, patient/family frustration, and non-harmful medical errors. The socioemotional route significantly impacted incidents, which had compassion as its starting point.

For the cognitive route, nursing professionals who had reservations about mental model sharing were more likely to experience miscommunication, particularly judgmental and conceptual failure types, when interacting with other medical staff. The former type involved errors in situational and information judgments and led to severe incidents, including adverse events and treatment/care delays. In contrast, the latter type, which represented insufficient information exchange and consensus, gave rise to milder incidents, such as patient/family frustration and non-harmful medical errors. This cognitive route was more prominent in interactions with medical staff than in those with patients. The rise of the patient safety movement in Japan was triggered by incidents such as fatalities due to deficiencies in handoffs during surgery.[21][22][23] In the current healthcare landscape, which emphasizes collaborative team-based care, effective mental model sharing and communication with other medical staff are essential for patient safety.

## GENERAL DISCUSSION

In Study 1, we interviewed experienced nurses with over 20 years of practical experience to explore their explanation skills. These nurses shared the nursing practice knowledge they had cultivated throughout their careers.[24] Their practical knowledge provided insights that effective explanatory communication in nursing practice required information transmission techniques and empathy toward the recipient. In assertive communication, respecting and accepting the other person’s opinions while asserting one’s own is essential. Similarly, accurate information transfer and an attitude that prioritizes the recipient’s feelings are required in explanatory nursing communication. Therefore, the two identified factors, socioemotional compassion, and cognitive mental model sharing, provide essential insights for nursing education and career development support. Specifically, to improve the quality of healthcare and patient safety, nurses’ training in communication skills should focus on fostering an attitude that goes beyond cognitive technicalities about information transmission and emphasizes consideration of the distressing position and feelings of the other person.

Seven identified communication sub-skills were common across patients/families and medical staff. Moreover, these sub-skills also demonstrated valid relationships with general communication skills. Therefore, although this study focused on nursing explanation skills, the identified skills and their structure were wider than nursing practice and applied to various acts of explaining to others. Safety science emphasizes the concept of “non-technical skills.”[25][26] In healthcare, non-technical skills (e.g., cognitive, social, and personal resource skills) related to diverse themes in psychology involving human subjects are necessary in addition to technical skills and knowledge.[27][28] Acquiring more general explanation skills is crucial for a nursing professional to excel with advanced nursing expertise. Regarding the characteristics of explanation skills, these factors were not defined by the type of actions such as “conveying,” “confirming,” or “questioning,” but rather by the purpose (e.g., explaining clearly and receiving in an empathetic manner). For example, the act of confirming included eight items, but these were distributed among “clarity” in mental model sharing (three items), “agreement” (two items), and “handover” (one item). In addition, compassion included two items related to “empathy.” This finding suggested that it is crucial to instruct students to consciously understand the purpose of their actions rather than just instructing them on behavioral levels (e.g., “let us confirm thoroughly”) in efforts to enhance explanatory communication skills in nursing education and career development support. In the example given earlier, confirmation is necessary for determining the other person’s level of understanding, aligning with the recipient’s expertise, and understanding the recipient’s emotions.

Nursing explanation skills were divided into two higher-order factors, and conveying information was distinctly cognitive. This study consistently provided insights into the importance of compassion in nursing explanations. Compassion has long been recognized as an essential element in nursing. In recent years, there has been an increasing demand for high-quality nursing, and the socioemotional importance of compassion has also grown.[29][30][31] However, it has been noted that compassion from hospital staff toward patients is often lacking.[32][33] Compassion is a fundamental concept in nursing, but evidence-based interventions and programs to enhance nurses’ compassion are scarce.[34] To improve the quality and safety of healthcare, education, and training for healthcare professionals, including nurses, the focus will need to shift toward compassion.

Notably, the negative impact of mental model sharing on patients and their families was not as strong as with medical staff. This insight suggested that healthcare professionals who have established a common “language” that includes medical terminology and styles (e.g., the “situation, background, evaluation, and recommendation” or SBAR technique that medical professionals use to conduct appropriate communication),[35] it is essential to employ both compassion and mental model sharing skills to ensure there are no discrepancies in information explanations. For patients and their families who often lack knowledge about medical matters and may be reluctant to accept inconvenient facts, nurses must be mindful of their feelings and prioritize compassion that considers the other’s perspective and emotions in explanations.[36]

## CONCLUSION

We developed a psychological scale to measure nursing explanation skills and elucidated their impact on incident occurrence through miscommunication. However, this study had limitations. One was the credibility of these insights. Therefore, further investigations with samples from various countries, regions, and hospitals are needed. Another was that the findings were from a survey and statistical analysis, representing only part of the process. Besides basic research, exploring how to apply these findings concurrently is imperative, as nurse attrition is a serious issue worldwide. Compassionate communication with patients, families, and other staff may foster positive relationships for nurses. Furthermore, accurate sharing of mental models can eliminate discrepancies and avoid conflicts. We hope to use these findings as evidence for practical considerations in supporting nursing professionals in building successful careers.

## Data Availability

All data produced in the present study are available upon reasonable request to the authors

## ACKNOWLEDGMENTS

We thank Edanz (https://jp.edanz.com/ac) for editing a draft of this manuscript.

## REFERENCES

1. The American Nurses Association. 2021. Nursing: Scope and Standards of Practice, 4th Edition.

2. The Florida Nurse Practice Act. 2023. Accessed Oct 13, 2023. http://www.leg.state.fl.us/Statutes/index.cfm?App_mode=Display_Statute&URL=0400-0499/0464/0464.html.

3. The National Commission for the Protection of Human Subjects of Biomedical and Behavioral Research. The Belmont Report: ethical principles and guidelines for protection of human subjects of biomedical and behavioral research. 1979. Accessed Oct 13, 2023. https://www.hhs.gov/ohrp/regulations-and-policy/belmont-report/read-the-belmont-report/index.html.

4. Umberfield E, et al. Using Incident Reports to Assess Communication Failures and Patient Outcomes. Jt Comm J Qual Patient Saf. 2019 Jun;45(6):406–413.

5. Alanazi FK, et al. Systematic review: Nurses’ safety attitudes and their impact on patient outcomes in acute-care hospitals. Nurs Open. 2022 Jan;9(1):30–43.

6. Lee A, et al. Root cause analysis of serious adverse events among older patients in the Veterans Health Administration. Jt Comm J Qual Patient Saf. 2014 Jun;40(6):253–62.

7. Humphrey KE, et al. Frequency and Nature of Communication and Handoff Failures in Medical Malpractice Claims. J Patient Saf. 2022 Mar 1;18(2):130–137.

8. Scott AM, et al; Project ACHIEVE Team. Understanding Facilitators and Barriers to Care Transitions: Insights from Project ACHIEVE Site Visits. Jt Comm J Qual Patient Saf. 2017 Sep;43(9):433–447.

9. Marrone M, et al. Consent and Complications in Health Care: The Italian Context. Healthcare (Basel). 2023 Jan 27;11(3):360.

10. Perrenoud B, et al. The effectiveness of health literacy interventions on the informed consent process of health care users: a systematic review protocol. JBI Database System Rev Implement Rep. 2015 Oct;13(10):82–94.

11. Gutiérrez-Puertas L, et al. Educational Interventions for Nursing Students to Develop Communication Skills with Patients: A Systematic Review. Int J Environ Res Public Health. 2020 Mar 26;17(7):2241.

12. Panczyk M, et al. Communication skills attitude scale: a translation and validation study in asample of registered nurses in Poland. BMJ Open. 2019 May 9;9(5):e028691.

13. Foronda C, et al. Interprofessional communication in healthcare: An integrative review. Nurse Educ Pract. 2016 Jul;19:36–40.

14. Fujimoto M, Daibo I. ENDCORE: A Hierarchical Structure Theory of Communication Skills. The Japanese Journal of Personality. 2007;15(3):347–361.

15. Fujimoto M. An Empirical and Conceptual Examination of the ENDCORE Model for Practical Work with Communication Skills. The Japanese Journal of Personality. 2013;22(2):156–167.

16. Blake RR, Mouton JS. The New Managerial Grid: Strategic New Insights into a Proven System for Increasing Organizational Productivity and Individual Effectiveness, Gulf Pub. 1979.

17. Jehn KA. A multimethod examination of the benefits and detriments of intragroup conflict. ASQ. 1995 Jun;42(2):256–282.

18. Green B, et al. Situational awareness - what it means for clinicians, its recognition and importance in patient safety. Oral Dis. 2017 Sep;23(6):721–725.

19. Edwards SJ, et al. Informed consent for clinical trials: in search of the “best” method. Soc Sci Med. 1998 Dec;47(11):1825–1840.

20. Neuhaus C, et al. Impact of a semi-structured briefing on the management of adverse events in anesthesiology: a randomized pilot study. BMC Anesthesiol. 2019 Dec 18;19(1):232.

21. Taneda K. Patient safety: History and recent updates in Japan. J. Natl. Inst. Public Health.2019;68(1):55–60.

22. Yokohama City University Hospital Accident Investigation Committee, Yokohama City University Hospital Accident Report. Accessed Oct 13, 2023.

23. https://www.yokohama-cu.ac.jp/kaikaku/bk2/bk21.html.

23. Committee for the Promotion of Measures to Prevent Medical Accidents at Tokyo Metropolitan Hospital: Report on Medical Accident at Tokyo Metropolitan Hiroo Hospital: Verification and Recommendations. 1999 Aug.

24. Benner P, et al. Clinical wisdom and interventions in critical care: A thinking-in-action approach. Philadelphia, W.B. Saunders.1999.

25. King HB, et al. TeamSTEPPS™: Team Strategies and Tools to Enhance Performance and Patient Safety. In: Henriksen K, Battles JB, Keyes MA, Grady ML, editors. Advances in Patient Safety: New Directions and Alternative Approaches (Vol. 3: Performance and Tools). Rockville (MD): Agency for Healthcare Research and Quality (US); 2008 Aug.

26. Flin R, et al. Safety at the sharp end: A guide to non-technical skills, Burlington, VA, Ashgate. 2008. 27.

27. Fujimoto M, et al. SAINTS: Self-Assessment Inventory of Non-Technical Skills for Medical Care. Healthcare and safety. 2021 Sep;13:36–43.

28. Fujimoto M, et al. Influence of Medical Staff’s Non-Technical Skills on Quality and Safety of Medical Care. Healthcare and safety. 2021 Sep;13:44–51.

29. Eriksson K. The alleviation of suffering--the idea of caring. Scand J Caring Sci. 1992;6(2):119–23.

30. Watson J. Nursing: the philosophy and science of caring. In Smith MC, et al., editors: Caring in Nursing Classics: An Essential Resource. Springer. 2008;243–264.

31. Dewar B, Cook F. Developing compassion through a relationship centred appreciative leadership programme. Nurse Educ Today. 2014 Sep;34(9):1258–1264.

32. Francis R. Independent Inquiry into Care Provided by Mid Staffordshire NHS Foundation Trust January 2005–March 2009. 2010. Accessed Oct 13, 2023. https://assets.publishing.service.gov.uk/government/uploads/system/uploads/attachment_data/file/279109/0375_i.pdf.

33. Francis R. Report of the Mid Staffordshire NHS Foundation Trust public inquiry. 2013 Feb. Accessed Oct 13, 2023. https://www.gov.uk/government/publications/report-of-the-mid-staffordshire-nhs-foundation-trust-public-inquiry.

34. Tehranineshat B, et al. Compassionate Care in Healthcare Systems: A Systematic Review. J Natl Med Assoc. 2019 Oct;111(5):546–554.

35. Müller M, et al. Impact of the communication and patient hand-off tool SBAR on patient safety: a systematic review. BMJ Open. 2018 Aug 23;8(8):e022202.

36. Seifart C, et al. Breaking bad news-what patients want and what they get: evaluating the SPIKES protocol in Germany. Ann Oncol. 2014 Mar;25(3):707–711.

